# Older Adults’ Attitudes Regarding COVID-19 and Associated Infection Control Measures in Shanghai, and Impact on Well-Being

**DOI:** 10.1101/2022.10.26.22281511

**Authors:** Lixian Cui, Zhiming Xu, Gabriela Lima de Melo Ghisi, Xia Liu, Sherry L Grace

## Abstract

This cross-sectional study investigated health management, well-being, and pandemic-related perspectives in Shanghainese adults ≥50 years during early and strict COVID-19 control measures. A self-report survey was administered via Wenjuanxing between March-April/2020. Items from the Somatic Symptom Scale, Patient Health Questionnaire-9 and Generalized Anxiety Disorder-7 were administered, as well as pandemic-specific questions. 1181 primarily married, retired females participated; Many had hypertension (n=521, 44.1%), coronary artery disease (CAD; n=201, 17.8%) and diabetes (n=171, 14.5%). While most respondents (n=868; 73.5%) were strictly following control measures (including limiting visits with children; n=390, 33.0%) and perceived they could tolerate that beyond 6 months (n=555;47.0%), they were optimistic about the future if control measures were continued (n=969;82.0%), and perceived impact would be temporary (n=646;64.7%). 52 of those with any condition (8.2%) and 19 of those without a condition (3.5%) reported the pandemic was impacting their health. Somatic symptoms were high (29.4±7.1/36), with Sleep & Cognitive symptoms highest. 24.4% and 18.9% of respondents had elevated depressive and anxious symptoms, respectively; greater distress was associated with lower income (p=0.018), having hypertension (p=0.001) and CAD (p<0.001), more negative perceptions of global COVID-19 control (p=0.004), COVID-19 spread (p≤0.001), impact on life and health (p<0.001), compliance with control measures (p<0.001), and shorter time control measures could be tolerated (p<0.001) in adjusted analyses. In the initial COVID-19 outbreak, most older adults were optimistic and resilient with regard to the epidemic and control measures. However, the distress of older adults is not trivial, particularly in those with health issues.

## INTRODUCTION

The COronaVIrus Disease (COVID-19) pandemic has resulted in major negative impacts for economies, health systems and citizens worldwide. While the impact of COVID-19 on the health of all people has been of major concern, particularly before the availability of vaccines or any treatment (Zheng et al., 2022), it has been a particular concern for those in dense urban areas where spread could be greater, as well as those at higher risk of complications. This includes older adults; For instance, hospitalization (i.e., 45%) and mortality rates (i.e., 80%) are much higher in individuals ≥65years (Shahid et al., 2020), as they often suffer from chronic conditions, many of which are themselves associated with increased risk (Izcovich et al., 2020).

Measures to control infection spread such as physical distancing often necessitate closure of essential businesses, such that individuals have well-founded anxiety about accessing food and medication, and stay-at-home orders mean they do not have access to places to be active or connect with others (Girum et al., 2021). There has been some study of the impacts of the pandemic and these associated control measures on the psychosocial well-being of older adults, such as anxiety and loneliness, suggesting variability, with certain subgroups at greater risk (De Pue et al., 2021; Jiang et al., 2020; Lee et al., 2020; Ping et al., 2020; Tian et al., 2020; Wong et al., 2020; Zhang et al., 2020; Zhao et al., 2020). This might be particularly true in China where filial piety is culturally integral, and control measures could impact this. Contrarily, there have also been some findings of a “well-being paradox”, such that at the beginning of the pandemic, communities rallied to support one another (and particularly older and vulnerable adults) and sense of well-being was bolstered (De Pue et al., 2021). It is also shown that older adults often have less psychological distress than their younger counterparts (Fiske et al., 2009).

Government policy varies worldwide in terms of implementation of control measures. Based on the COVID-19 stringency index, such strategies are among the most stringent in China (Hale et al., 2021). For example, in one of the world’s most populous cities of Shanghai, policies enforced included no gatherings of more than 10 people (big events such as wedding and birthday parties were postponed), not being able to leave Shanghai with visitations being restricted, non-residents of the community not allowed to enter, and some specialist outpatient services cancelled. Therefore, the objective of this study was to assess the impact of the pandemic on older adults, in the populous city of Shanghai, during the first wave --when most cases were in China, there was much uncertainty and no vaccine or treatments were available.

Impacts on: (1) psychosocial well-being, and (2) perceptions related to the pandemic and its’ control were investigated. Vulnerability and protective factors for elevated depressive and anxiety symptoms were also explored.

## METHODS

### Design and Procedure

Ethics approval was secured from the Ethics Committee of Xinhua Hospital, affiliated with Shanghai Jiaotong University School of Medicine (XHEC-D-2022-179). The voluntary and anonymous online survey was created by the Cardiac Rehabilitation Group of the Health Risk Assessment and Control Committee of the Chinese Preventive Medical Association in Mandarin. Data collection for this cross-sectional study was undertaken between March and April in 2020. The survey was disseminated via the web-based survey platform Wenjuanxing through WeChat by the Community Doctor Group including 36 community doctors from all the 16 districts in Shanghai, which belongs to the Shanghai Association of Chinese Integrative Medicine.

### Setting and Participants

Participants of this study were adults ≥50 years old residing in a community for seniors in Shanghai. Exclusion criteria were the following: severe cognitive impairments or any conditions that prevented respondents from being able to understand and voluntarily agree to participate in the research.

### Measures

All items were self-reported. Non-psychometrically validated items were generated by the Cardiac Rehabilitation group (see above), and respondents were asked to respond based on their experience in the prior two weeks.

The Patient Health Questionnaire-9 (PHQ-9) was administered (Kroenke et al., 2001). The PHQ-9 assesses depression symptom severity in alignment with diagnostic criteria for a major depressive disorder. For each item, the patient is asked to rate symptoms over the last 2 weeks, with each item rated on a 4-point Likert scale from 0 (“not at all”) to 3 (“almost every day”). Total scale scores range from 0 to 27, with higher scores indicating greater severity of depression. The cut-off points are 5, 10 and 15, indicating mild, moderate, and severe depression, respectively. PHQ-9 is considered valid and reliable among Chinese people (Wang et al., 2014).

The Generalized Anxiety Disorder (GAD-7) questionnaire was administered to evaluate symptom of anxiety. It comprises seven items, with each item scored on a 4-point Likert scale ranging from 0 (“not at all”) to 3 (“almost every day”). Total scale scores range from 0 to 21, with higher scores indicating greater anxiety. Cut-off points of 5, 10, and 15 represent mild, moderate, and severe levels of anxiety, respectively (Lowe et al., 2008). The GAD-7 has been proven to have good reliability and validity in Chinese populations (He et al., 2010).

The Chinese version of the Somatic Self-rating Scale (SSS) was also administered to assess physical (i.e., half the items query each body system), depressive, and anxiety symptoms as well as sleep and cognitive issues (Zhuang et al., 2010). Thus, the SSS is used to measure not only somatic symptoms, but also psychosocial well-being. It consists of 20 items categorized in four domains, with each item is rated on a 4-point scale. The total score ranges from 20 to 80, with scores ranging from 20 to 29, 30–39, 40–59 and≥60 correspond to normal, mild, moderate and severe somatic symptom disorder respectively (Wu et al., 2022). The SSS has been proven to have good psychometric properties, including test–retest reliability of 0.9 and Cronbach’s α of 0.89. In addition, the correlation coefficient between the total score of SSS and the self-rating anxiety scale (SAS) and the self-rating depression scale (SDS) was 0.80 and 0.74 respectively (Zhuang et al., 2010).

### Statistical Analyses

All data analysis was performed using IBM SPSS statistics version 25.0, with p<0.05 considered statistically significant. Descriptive analysis was first performed. The associations of physical and mental health with perceived pandemic impact on their health were tested using chi-square and t-tests, as applicable.

The association of pandemic-related attitudes and health impact with depressive and anxious symptoms were assessed using t-test and chi-square, as applicable. Finally, multivariate analysis of variance assessing significant correlates of depressive and anxious symptoms based on the aforementioned analyses as well as associated sociodemographic and clinical correlates were tested using multivariate analysis of variance (MANOVA).

## RESULTS

Of 1181 responding participants, most were female, over 65 years, married and living with their partner only, had moderate education, and were retired with the associated healthcare coverage (Table 1). As shown in Table 2, almost half of participants had hypertension, almost one-in-five had coronary artery disease, with another 15% diabetes; 546 (46.2%) had no assessed condition.

**Table 1:**
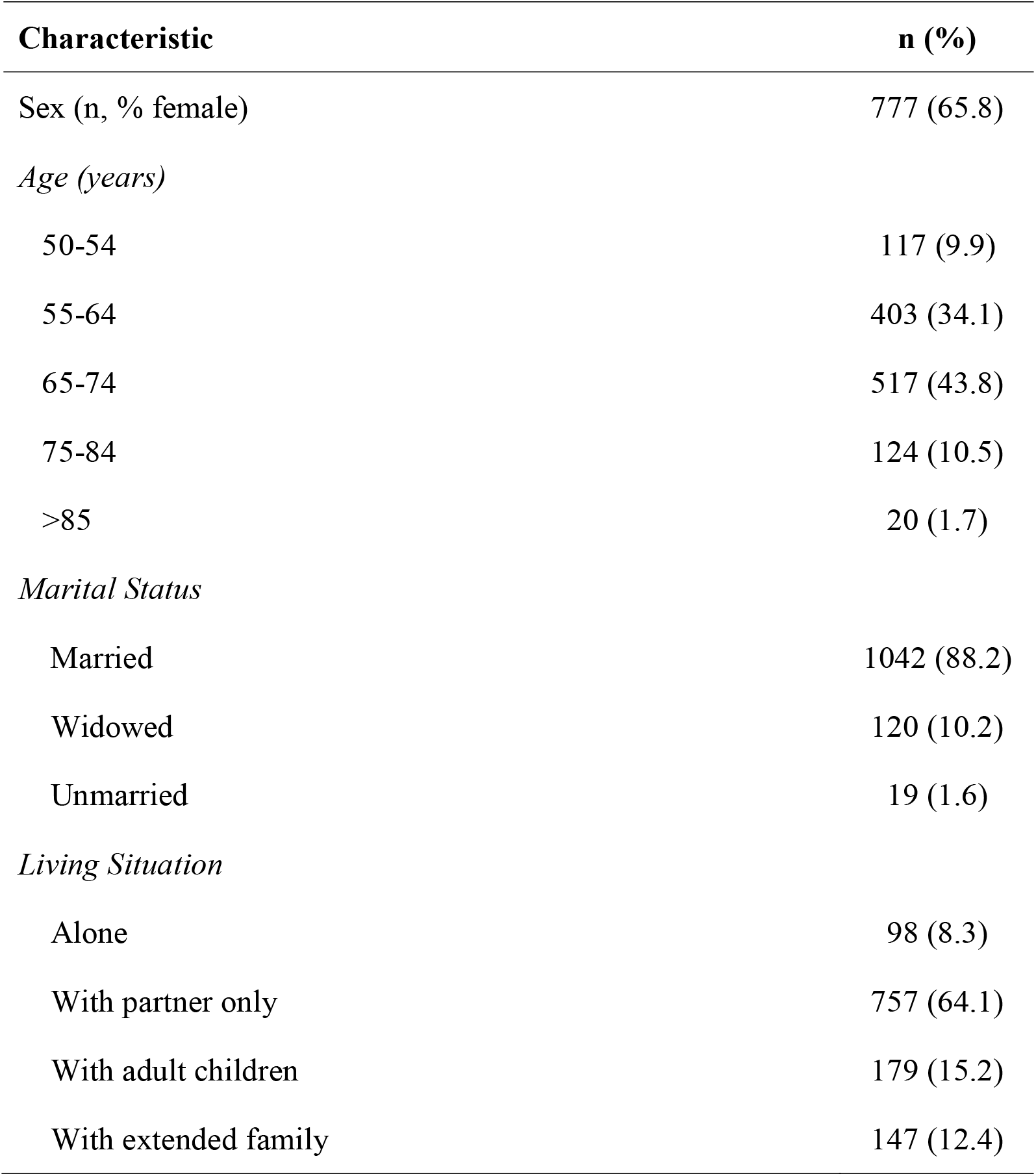
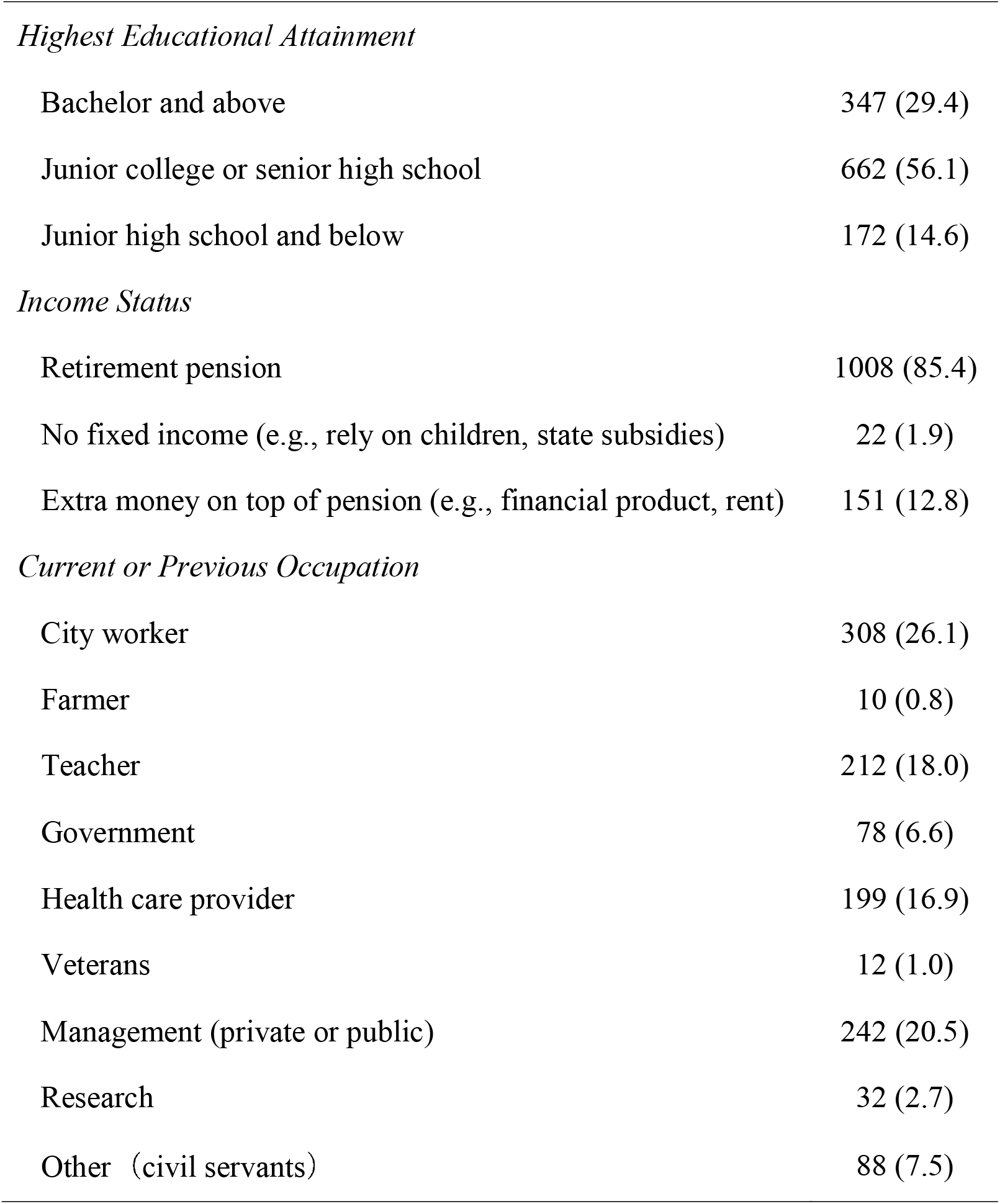
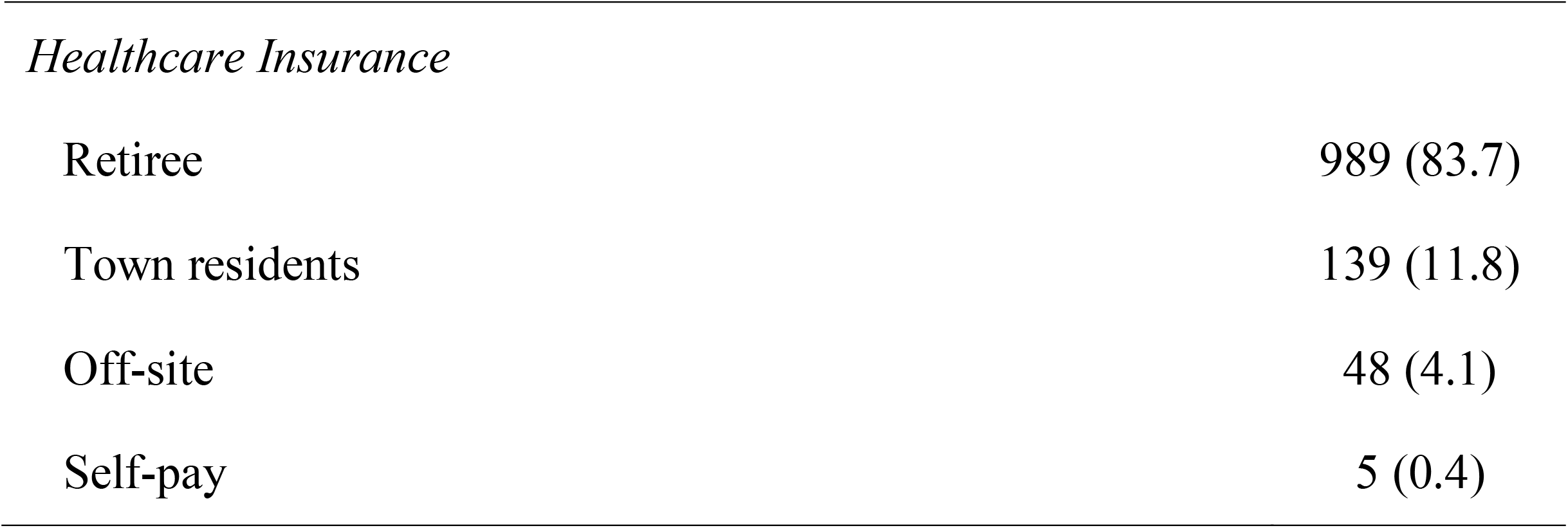
Participant Sociodemographic Characteristics, N=1181

| Characteristic | n (%) |
| --- | --- |
| Sex (n, % female) | 777 (65.8) |
| *Age (years)* |  |
| 50-54 | 117 (9.9) |
| 55-64 | 403 (34.1) |
| 65-74 | 517 (43.8) |
| 75-84 | 124 (10.5) |
| >85 | 20 (1.7) |
| *Marital Status* |  |
| Married | 1042 (88.2) |
| Widowed | 120 (10.2) |
| Unmarried | 19 (1.6) |
| *Living Situation* |  |
| Alone | 98 (8.3) |
| With partner only | 757 (64.1) |
| With adult children | 179 (15.2) |
| With extended family | 147 (12.4) |
| *Highest Educational Attainment* |  |
| Bachelor and above | 347 (29.4) |
| Junior college or senior high school | 662 (56.1) |
| Junior high school and below | 172 (14.6) |
| *Income Status* |  |
| Retirement pension | 1008 (85.4) |
| No fixed income (e.g., rely on children, state subsidies) | 22 (1.9) |
| Extra money on top of pension (e.g., financial product, rent) | 151 (12.8) |
| *Current or Previous Occupation* |  |
| City worker | 308 (26.1) |
| Farmer | 10 (0.8) |
| Teacher | 212 (18.0) |
| Government | 78 (6.6) |
| Health care provider | 199 (16.9) |
| Veterans | 12 (1.0) |
| Management (private or public) | 242 (20.5) |
| Research | 32 (2.7) |
| Other (civil servants) | 88 (7.5) |
| *Healthcare Insurance* |  |
| Retiree | 989 (83.7) |
| Town residents | 139 (11.8) |
| Off-site | 48 (4.1) |
| Self-pay | 5 (0.4) |

**Table 2:** Participant Physical and Mental Health and Association with Perceived Pandemic Impact on Health, N=1181

|  | n (%) /<br>mean ± SD | Perceived Impact of Pandemic on Health Condition |  |  |  |
| --- | --- | --- | --- | --- | --- |
|  |  | No Effect<br>(n=1110) | Effect<br>(n=71) | X²/t | P |
| *Chronic Condition* |  |  |  |  |  |
| Hypertension | 521 (44.1) | 480 (43.2) | 41 (57.7) | 5.694 | 0.017 |
| Coronary artery disease | 210 (17.8) | 189 (17.0) | 21 (29.6) | 7.190 | 0.007 |
| Diabetes | 171 (14.5) | 156 (14.1) | 15 (21.1) | 2.696 | 0.101 |
| Cerebrovascular disease | 83 (7.0) | 68 (6.1) | 15 (21.1) | 22.981 | <0.001 |
| Renal disease | 29 (2.5) | 27 (2.4) | 2 (2.8) | 0.041 | 0.839 |
| Heart failure | 44 (3.7) | 32 (2.9) | 12 (16.9) | 32.758 | <0.001 |
| None of the above | 546 (46.2) | 527 (47.5) | 19 (26.8) | 11.522 | 0.001 |
| One or more of above chronic conditions | 635 (53.8) | 583 (52.5) | 52 (73.2) |  |  |
| *Mental Health* |  |  |  |  |  |
| History psychiatric disorder (n, % yes) | 62 (5.2) | 52 (4.7) | 10 (14.1) | 10.039 | 0.002 |
| Taking psychoactive medication (n, % yes) | 61 (5.2) | 51 (4.6) | 10 (14.1) | 10.408 | 0.001 |
| Major Life Event (n, % yes) | 133 (11.3) | 122 (11.0) | 11 (15.5) | 1.353 | 0.245 |
| Somatic Self-rating Scale -Total (/80) | 29.40 ± 7.11 | 29.01 ± 6.71 | 35.58 ± 9.85 | 5.540 | <0.001 |
| Somatic Symptoms (/36) | 13.32 ± 3.35 | 13.17 ± 3.24 | 15.72 ± 4.11 | 5.117 | <0.001 |
| Anxiety symptoms (/20) | 6.89 ± 1.90 | 6.79 ± 1.78 | 8.51 ± 2.82 | 5.073 | <0.001 |
| Depressive symptoms (/16) | 5.85 ± 1.77 | 5.76 ± 1.67 | 7.30 ± 2.55 | 5.019 | <0.001 |
| Sleep & cognitive symptoms (/8) | 3.33 ± 1.09 | 3.29 ± 1.04 | 4.06 ± 1.48 | 4.299 | <0.001 |
| SSS normal (20-29) | 681(57.7) | 658 (59.3) | 23 (32.4) | 37.848 | <0.001 |
| SSS mild (30-39) | 409 (34.6) | 381 (34.3) | 28 (39.4) |  |  |
| SSS moderate (40-59) | 87 (7.4) | 68 (6.1) | 19 (26.8) |  |  |
| SSS severe (≥60) | 4 (0.3) | 3 (0.3) | 1 (1.4) |  |  |
| SSS elevated ( > 36) | 185 (15.7) | 154 (13.9) | 31 (43.7) | 44.821 | <0.001 |
| Depressive Symptoms (PHQ-9) | 2.50 ± 3.15 | 2.29 ± 2.81 | 5.79 ± 5.52 | 5.295 | <0.001 |
| Elevated (5-27) | 238 (20.2) | 203 (18.3) | 35 (49.3) | 39.873 | <0.001 |
| Sub-clinical (0-4) | 943 (79.8) | 907 (81.7) | 36 (50.7) |  |  |
| Anxiety Symptoms (GAD-7) | 1.88 ± 3.04 | 1.68 ± 2.71 | 4.94 ± 5.42 | 5.033 | <0.001 |
| Elevated (5-21) | 201 (17.0) | 168 (15.1) | 33 (46.5) | 46.421 | <0.001 |
| Sub-clinical (0-4) | 980 (83.0) | 942 (84.9) | 38 (53.5) |  |  |

SD, standard deviation; GAD-7, Generalized Anxiety Disorder; PHQ-9, The Patient Health Questionnaire-9; SSS, Somatic Self-rating Scale.

As also shown in Table 2, 62 (5%) reported a psychiatric diagnosis. Of those reporting taking a psychoactive medication, these included: sleeping pills (n=28; 45.9%), anti-depressants (n=42; 68.9%) or both (n=9; 14.8%). Of those reporting a pre-pandemic major life event, these included: death of a family member (n=14; 53.8%), critical illness of family members or self (n=11; 42.3%) and economic pressure (n=1; 3.8%).

### Impact of Pandemic

Over two-thirds of participants perceived the impact of the pandemic on their life would be temporary (Table 3). Almost three-quarters strictly complied with COVID-19 prevention and control measures. Almost half of participants reported they could tolerate strict control measures for a long time, with over 20% reporting they could endure it for 6 months or 3 months. Not considering those participants who were living with their children, most isolated from their children, having no visits with them. Almost three-quarters transitioned to exercise in their home, and only 8.2% reported the pandemic was having an impact on their health.

**Table 3:**
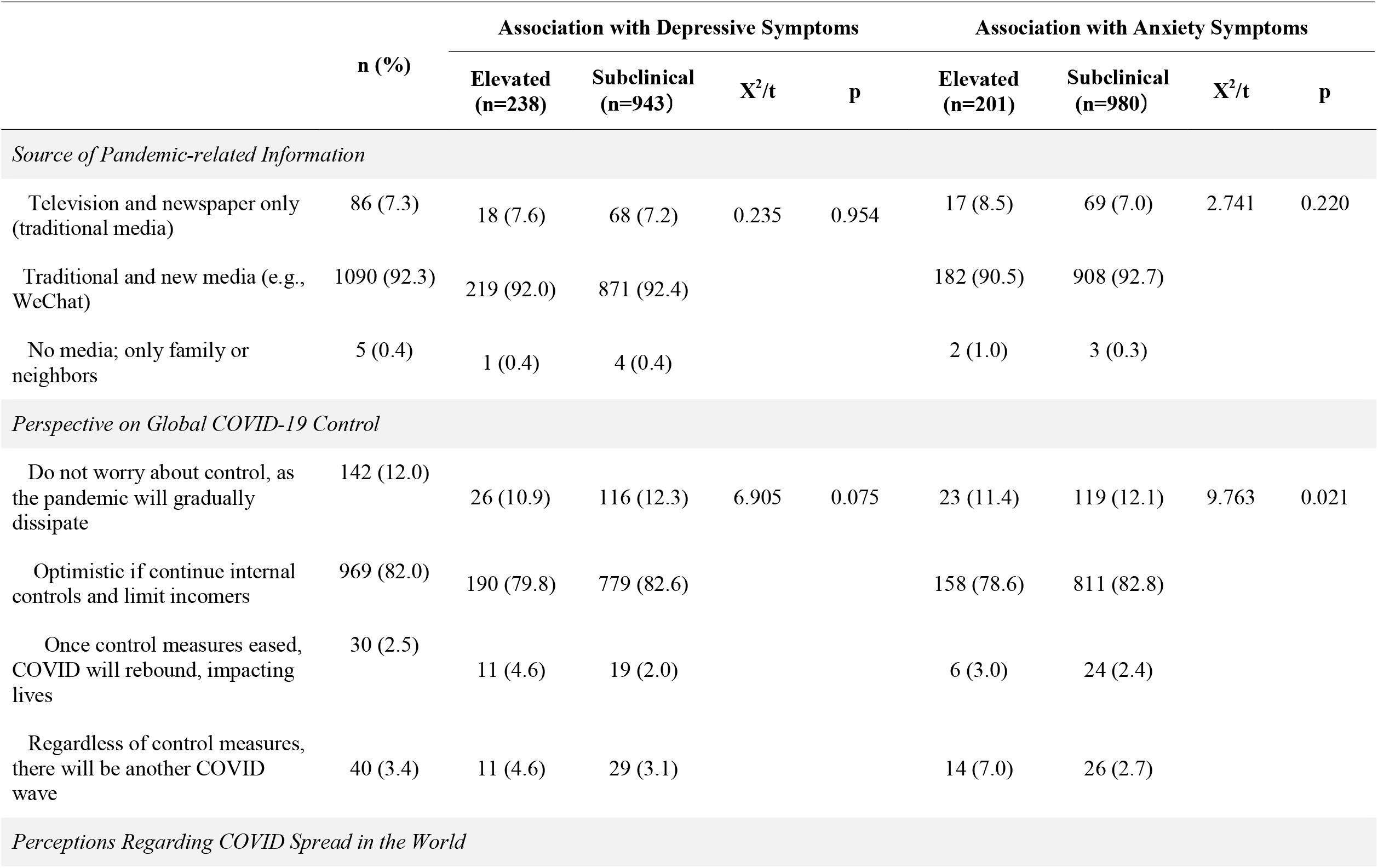
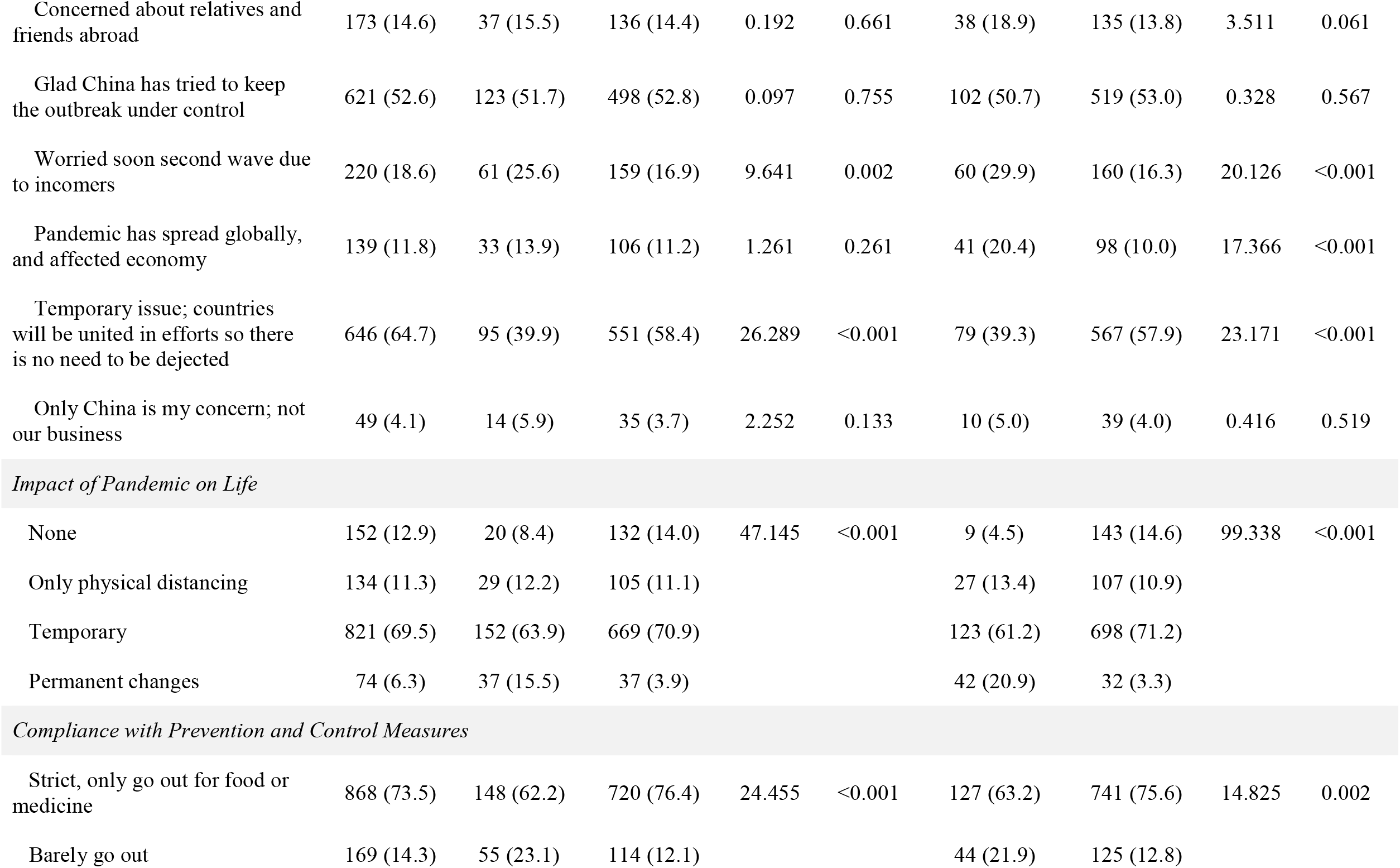
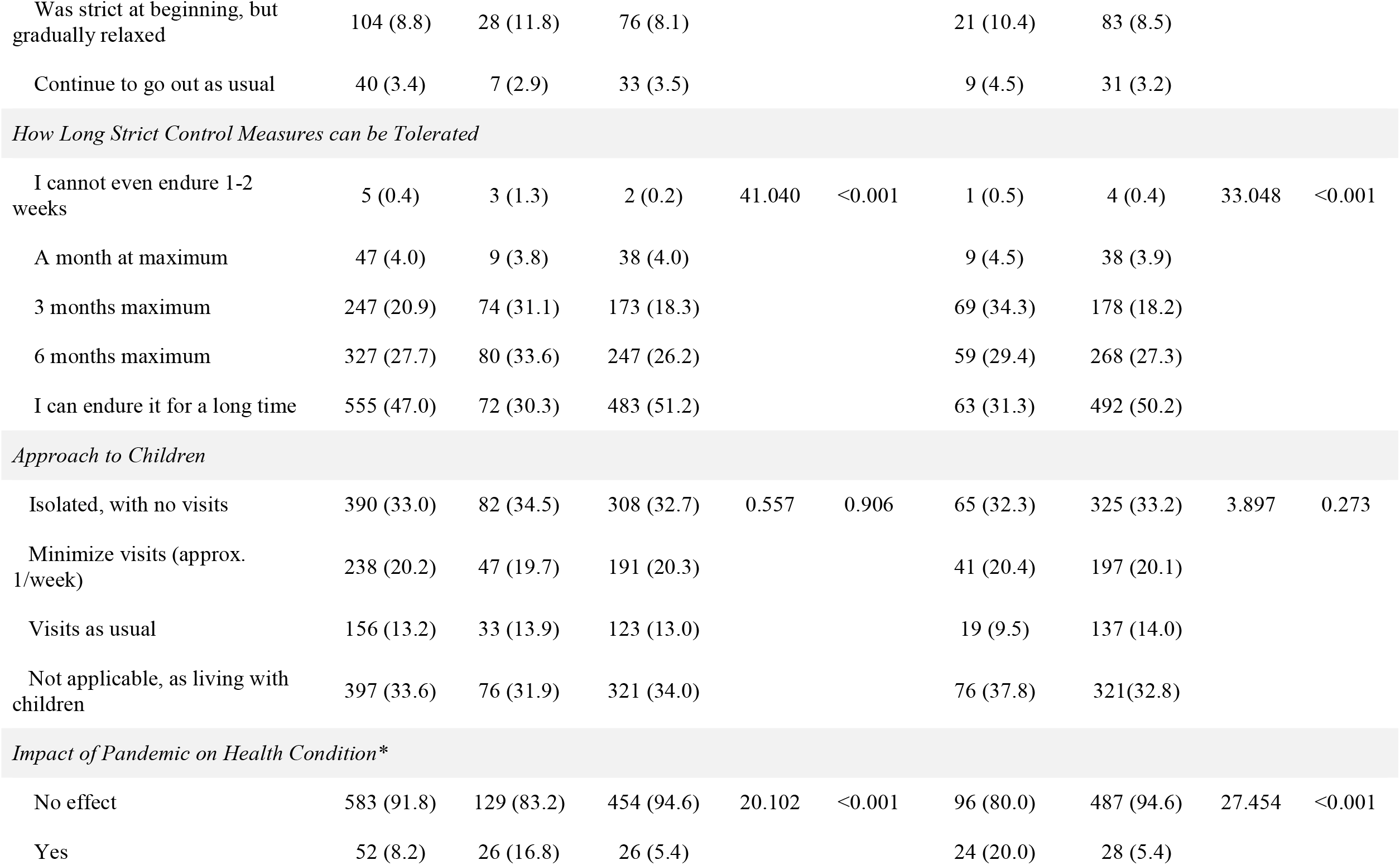
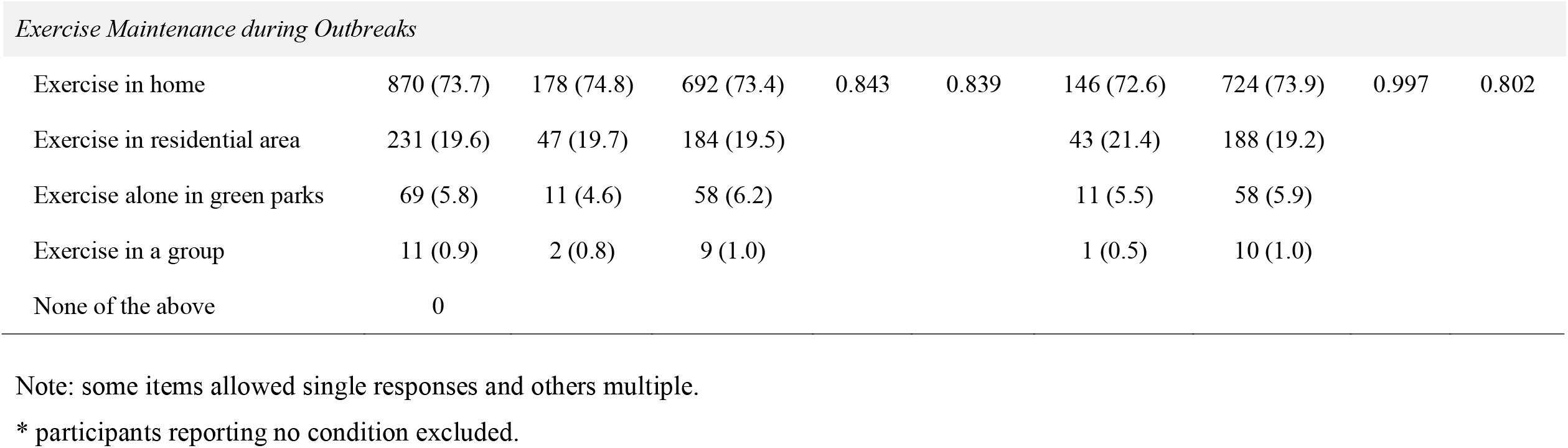
Pandemic Attitudes and Health Impact, and their Associations with Depressive and Anxious Symptoms, N=1181

|  | n (%) | Association with Depressive Symptoms |  |  |  | Association with Anxiety Symptoms |  |  |  |
| --- | --- | --- | --- | --- | --- | --- | --- | --- | --- |
|  |  | Elevated<br>(n=238) | Subclinical<br>(n=943) | X<sup>2</sup>/t | p | Elevated<br>(n=201) | Subclinical<br>(n=980) | X<sup>2</sup>/t | p |
| Source of Pandemic-related Information |  |  |  |  |  |  |  |  |  |
| Television and newspaper only<br>(traditional media) | 86 (7.3) | 18 (7.6) | 68 (7.2) | 0.235 | 0.954 | 17 (8.5) | 69 (7.0) | 2.741 | 0.220 |
| Traditional and new media (e.g.,<br>WeChat) | 1090 (92.3) | 219 (92.0) | 871 (92.4) |  |  | 182 (90.5) | 908 (92.7) |  |  |
| No media; only family or<br>neighbors | 5 (0.4) | 1 (0.4) | 4 (0.4) |  |  | 2 (1.0) | 3 (0.3) |  |  |
| Perspective on Global COVID-19 Control |  |  |  |  |  |  |  |  |  |
| Do not worry about control, as<br>the pandemic will gradually<br>dissipate | 142 (12.0) | 26 (10.9) | 116 (12.3) | 6.905 | 0.075 | 23 (11.4) | 119 (12.1) | 9.763 | 0.021 |
| Optimistic if continue internal<br>controls and limit incomers | 969 (82.0) | 190 (79.8) | 779 (82.6) |  |  | 158 (78.6) | 811 (82.8) |  |  |
| Once control measures eased,<br>COVID will rebound, impacting<br>lives | 30 (2.5) | 11 (4.6) | 19 (2.0) |  |  | 6 (3.0) | 24 (2.4) |  |  |
| Regardless of control measures,<br>there will be another COVID<br>wave | 40 (3.4) | 11 (4.6) | 29 (3.1) |  |  | 14 (7.0) | 26 (2.7) |  |  |
| Perceptions Regarding COVID Spread in the World |  |  |  |  |  |  |  |  |  |
| Concerned about relatives and friends abroad | 173 (14.6) | 37 (15.5) | 136 (14.4) | 0.192 | 0.661 | 38 (18.9) | 135 (13.8) | 3.511 | 0.061 |
| Glad China has tried to keep the outbreak under control | 621 (52.6) | 123 (51.7) | 498 (52.8) | 0.097 | 0.755 | 102 (50.7) | 519 (53.0) | 0.328 | 0.567 |
| Worried soon second wave due to incomers | 220 (18.6) | 61 (25.6) | 159 (16.9) | 9.641 | 0.002 | 60 (29.9) | 160 (16.3) | 20.126 | <0.001 |
| Pandemic has spread globally, and affected economy | 139 (11.8) | 33 (13.9) | 106 (11.2) | 1.261 | 0.261 | 41 (20.4) | 98 (10.0) | 17.366 | <0.001 |
| Temporary issue; countries will be united in efforts so there is no need to be dejected | 646 (64.7) | 95 (39.9) | 551 (58.4) | 26.289 | <0.001 | 79 (39.3) | 567 (57.9) | 23.171 | <0.001 |
| Only China is my concern; not our business | 49 (4.1) | 14 (5.9) | 35 (3.7) | 2.252 | 0.133 | 10 (5.0) | 39 (4.0) | 0.416 | 0.519 |
| *Impact of Pandemic on Life* |  |  |  |  |  |  |  |  |  |
| None | 152 (12.9) | 20 (8.4) | 132 (14.0) | 47.145 | <0.001 | 9 (4.5) | 143 (14.6) | 99.338 | <0.001 |
| Only physical distancing | 134 (11.3) | 29 (12.2) | 105 (11.1) |  |  | 27 (13.4) | 107 (10.9) |  |  |
| Temporary | 821 (69.5) | 152 (63.9) | 669 (70.9) |  |  | 123 (61.2) | 698 (71.2) |  |  |
| Permanent changes | 74 (6.3) | 37 (15.5) | 37 (3.9) |  |  | 42 (20.9) | 32 (3.3) |  |  |
| *Compliance with Prevention and Control Measures* |  |  |  |  |  |  |  |  |  |
| Strict, only go out for food or medicine | 868 (73.5) | 148 (62.2) | 720 (76.4) | 24.455 | <0.001 | 127 (63.2) | 741 (75.6) | 14.825 | 0.002 |
| Barely go out | 169 (14.3) | 55 (23.1) | 114 (12.1) |  |  | 44 (21.9) | 125 (12.8) |  |  |
| Was strict at beginning, but gradually relaxed | 104 (8.8) | 28 (11.8) | 76 (8.1) |  |  | 21 (10.4) | 83 (8.5) |  |  |
| Continue to go out as usual | 40 (3.4) | 7 (2.9) | 33 (3.5) |  |  | 9 (4.5) | 31 (3.2) |  |  |
| *How Long Strict Control Measures can be Tolerated* |  |  |  |  |  |  |  |  |  |
| I cannot even endure 1-2 weeks | 5 (0.4) | 3 (1.3) | 2 (0.2) | 41.040 | <0.001 | 1 (0.5) | 4 (0.4) | 33.048 | <0.001 |
| A month at maximum | 47 (4.0) | 9 (3.8) | 38 (4.0) |  |  | 9 (4.5) | 38 (3.9) |  |  |
| 3 months maximum | 247 (20.9) | 74 (31.1) | 173 (18.3) |  |  | 69 (34.3) | 178 (18.2) |  |  |
| 6 months maximum | 327 (27.7) | 80 (33.6) | 247 (26.2) |  |  | 59 (29.4) | 268 (27.3) |  |  |
| I can endure it for a long time | 555 (47.0) | 72 (30.3) | 483 (51.2) |  |  | 63 (31.3) | 492 (50.2) |  |  |
| *Approach to Children* |  |  |  |  |  |  |  |  |  |
| Isolated, with no visits | 390 (33.0) | 82 (34.5) | 308 (32.7) | 0.557 | 0.906 | 65 (32.3) | 325 (33.2) | 3.897 | 0.273 |
| Minimize visits (approx. 1/week) | 238 (20.2) | 47 (19.7) | 191 (20.3) |  |  | 41 (20.4) | 197 (20.1) |  |  |
| Visits as usual | 156 (13.2) | 33 (13.9) | 123 (13.0) |  |  | 19 (9.5) | 137 (14.0) |  |  |
| Not applicable, as living with children | 397 (33.6) | 76 (31.9) | 321 (34.0) |  |  | 76 (37.8) | 321 (32.8) |  |  |
| *Impact of Pandemic on Health Condition\** |  |  |  |  |  |  |  |  |  |
| No effect | 583 (91.8) | 129 (83.2) | 454 (94.6) | 20.102 | <0.001 | 96 (80.0) | 487 (94.6) | 27.454 | <0.001 |
| Yes | 52 (8.2) | 26 (16.8) | 26 (5.4) |  |  | 24 (20.0) | 28 (5.4) |  |  |
| *Exercise Maintenance during Outbreaks* |  |  |  |  |  |  |  |  |  |
| Exercise in home | 870 (73.7) | 178 (74.8) | 692 (73.4) | 0.843 | 0.839 | 146 (72.6) | 724 (73.9) | 0.997 | 0.802 |
| Exercise in residential area | 231 (19.6) | 47 (19.7) | 184 (19.5) |  |  | 43 (21.4) | 188 (19.2) |  |  |
| Exercise alone in green parks | 69 (5.8) | 11 (4.6) | 58 (6.2) |  |  | 11 (5.5) | 58 (5.9) |  |  |
| Exercise in a group | 11 (0.9) | 2 (0.8) | 9 (1.0) |  |  | 1 (0.5) | 10 (1.0) |  |  |
| None of the above | 0 |  |  |  |  |  |  |  |  |

Note: some items allowed single responses and others multiple.
\* participants reporting no condition excluded.

Psychosocial well-being early during the pandemic was compromised (Table 2). Mean SSS scores revealed approximately 42.3% of participants had elevated symptoms; Impacts on sleep and cognition were mild. Approximately 20% of participants had elevated depressive symptoms, and the same had elevated anxiety.

As shown in Table 2, participants with hypertension, coronary artery disease, cerebrovascular disease and heart failure perceived an impact of the pandemic on their health whereas participants without these conditions did not. Those perceiving an impact of the pandemic on their health condition also were significantly more likely to have a history of a psychiatric disorder, and correspondingly to have significantly greater somatic symptoms (all subscales) as well as depressive and anxiety symptoms.

As shown in Table 3, participants with elevated depressive symptoms were more worried about future waves and that the pandemic was not temporary, were more likely to barely go out, to be unable to endure strict control measures over prolonged periods and perceived significantly greater impact of the pandemic on their health when compared to participants with subclinical symptoms. Participants with elevated anxiety symptoms were significantly less optimistic about global control of the pandemic, were more worried about future waves due to incomers and the economic impact globally of the pandemic, were less likely to view the pandemic spread and impacts as temporary, were less likely to go out and reported they could endure strict control measures for a shorter time than participants with subclinical symptoms (Table 3).

Table 4 demonstrates that, in an adjusted MANOVA model, the following correlates were associated with greater depressive and anxiety symptoms: lower income, having hypertension, and coronary artery disease, negative perceptions regarding global COVID-19 control, some more negative perceptions regarding COVID-19 spread, of the impact of pandemic on their life, greater compliance with prevention and control measures, perceive negative impact of the pandemic on health condition and were less able to tolerate strict COVID-19 control measures over time.

**Table 4:** Multivariate Analysis of Variance Assessing Correlates of Depressive and Anxious Symptoms

| | Pillai's Trace | Wilk's $\lambda$ | F | P | $\eta^2$ |
| --- | --- | --- | --- | --- | --- |
| Sex | 0.002 | 0.998 | 1.245 | 0.288 | 0.002 |
| Age | 0.011 | 0.989 | 1.589 | 0.123 | 0.005 |
| Income Status | 0.010 | 0.990 | 2.973 | 0.018 | 0.005 |
| Hypertension | 0.012 | 0.988 | 7.071 | 0.001 | 0.012 |
| Coronary Artery Disease | 0.034 | 0.966 | 20.573 | <0.001 | 0.034 |
| Perspective on Global COVID-19 control | 0.016 | 0.984 | 3.224 | 0.004 | 0.008 |
| *Perceptions Regarding COVID-19 Spread* |  |  |  |  |  |
| Concerned about relatives and friends abroad | 0.011 | 0.989 | 6.826 | 0.001 | 0.011 |
| Glad China has tried to keep the outbreak under control | 0.003 | 0.997 | 1.896 | 0.151 | 0.003 |
| Worried soon second wave due to incomers | 0.022 | 0.978 | 13.151 | <0.001 | 0.022 |
| Pandemic has spread globally, and affected economy | 0.022 | 0.978 | 13.295 | <0.001 | 0.022 |
| Temporary issue; countries will be united in efforts so there is no need to be dejected | 0.029 | 0.971 | 17.368 | <0.001 | 0.029 |
| Only China is my concern; not our business | 0.002 | 0.998 | 1.288 | 0.276 | 0.002 |
| Impact of Pandemic on Life | 0.101 | 0.899 | 20.953 | <0.001 | 0.051 |
| Compliance with Prevention and Control Measures | 0.026 | 0.974 | 5.253 | <0.001 | 0.013 |
| Impact of Pandemic on Health Condition | 0.077 | 0.923 | 49.230 | <0.001 | 0.077 |
| How Long Strict Control Measures can be Tolerated | 0.052 | 0.948 | 7.824 | <0.001 | 0.026 |

## DISCUSSION

This study examined the impact of the COVID-19 pandemic and its’ associated control measures in a large sample of older adults, over half of whom had a chronic condition. It was undertaken in the context of some of the most stringent control measures globally in one of the most populous cities in the world (World of Atlas, 2022), early in the pandemic when uncertainty was high and there were no known treatments or vaccines. While some studies have examined this in the context of “western” cultures where older adults are more isolated and may live in retirement facilities, the context in China is different, with most older adults living with their spouses. Nevertheless, consistent with findings in other countries (Lorant et al., 2021; McGinty et al., 2022; Sams et al., 2021; Xiong et al., 2020), results show elevated distress, particularly when compared to the general population or medical outpatients more broadly (e.g., 5-10% depressive symptoms) (Bradley and Rumsfeld, 2015).

Indeed, while approximately one-quarter of older adults had elevated depressive symptoms and one-fifth anxiety, and most were only going out for food or medicine and limiting visits with children, there was still much optimism and faith the impacts were temporary and resignation to tolerate long-term control measures. According to Jiang et al. (2020), older Chinese adults were less worried about COVID-19 compared to younger Chinese adults during the initial outbreak; however, as the disease spread, their worries gradually increased. Studies by Tian et al. (2020), Wong et al. (2020) and Zhang et al. (2020) reiterate the negative psychological impact of COVID-19 on Chinese adults.

Study results are particularly concerning given the link between psychosocial well-being and chronic disease onset and outcomes (Lichtman et al., 2014). There are effective treatments for sleep issues (Ye et al., 2016), depression (Wilson et al., 2001) and anxiety (Hunot et al., 2007), although means to bolster support are less well-established (Fakoya et al., 2020).

Research into internet-based delivery of evidence-based therapies has been increasing and supportive, although some older adults do not have access to the technology or the digital literacy to take advantage of this COVID-friendly modality. Effective pharmacotherapy does not exist for all these conditions, and particularly sleep and anti-anxiety medications can be more hazardous in older adults (Schroeck et al., 2016).

There are some directions for future research stemming from this study. Whether patients or close contacts had COVID-19 during the period of study was not considered or had to quarantine; more research is needed. Other studies have shown impact on mobility (Beauchamp et al., 2022), which were not considered herein. Exercise was assessed herein, with most older adults reporting they were exercising at home, but given more objective reports (Castañeda-Babarro et al., 2020) it is likely activity was insufficient for optimal primary or secondary prevention (Santiago de Araújo Pio et al., 2017), thus ultimately leading to likely mobility issues. Moreover, research on the impacts of the pandemic and control measures over time is warranted, to determine how older adults cope in the longer-term (Robinson et al., 2022).

Caution is warranted when interpreting these results. First, this is a cross-sectional study, so the design precludes causal or directional determinations, and we cannot test how psychosocial well-being differs from pre-pandemic levels. Second, the results may not be generalizable beyond China with its specific political and cultural context and the city of Shanghai more specifically, and where the stringency of control measures was high.

Nevertheless, the population of Shanghai is very large, and findings can inform other governments when considering their control measures. Moreover, similarity of the sample to the larger population is not known given the recruitment strategy, designed to quickly secure data during the lockdown, but where full reach and non-responders were not collated. Third, all data were self-report, and hence there may have been socially desirable responding or other measurement error, and conditions were not verified through structured clinical interview or medical charts. Fourth, multiple comparisons were performed, inflating potential type 1 error.

In conclusion, in this sample of older adults in Shanghai at the beginning of the pandemic, half of whom had a chronic condition and were strictly following prevention and control measures, while optimism was high, levels of somatic symptoms as well as depressive and anxiety symptoms were higher than normative levels. Ensuring access to evidence-based treatment via technology in those who have the digital literacy would have been key at the beginning of the pandemic, knowing now what we do about its’ duration. More research on the longer-term physical and psychosocial impacts of COVID-19 and the associated prevention and control measures on older adults is needed.

## Data Availability

All data produced in the present work are contained in the manuscript

## Acknowledgements

none

## FIGURE LEGEND

**Figure 1:**
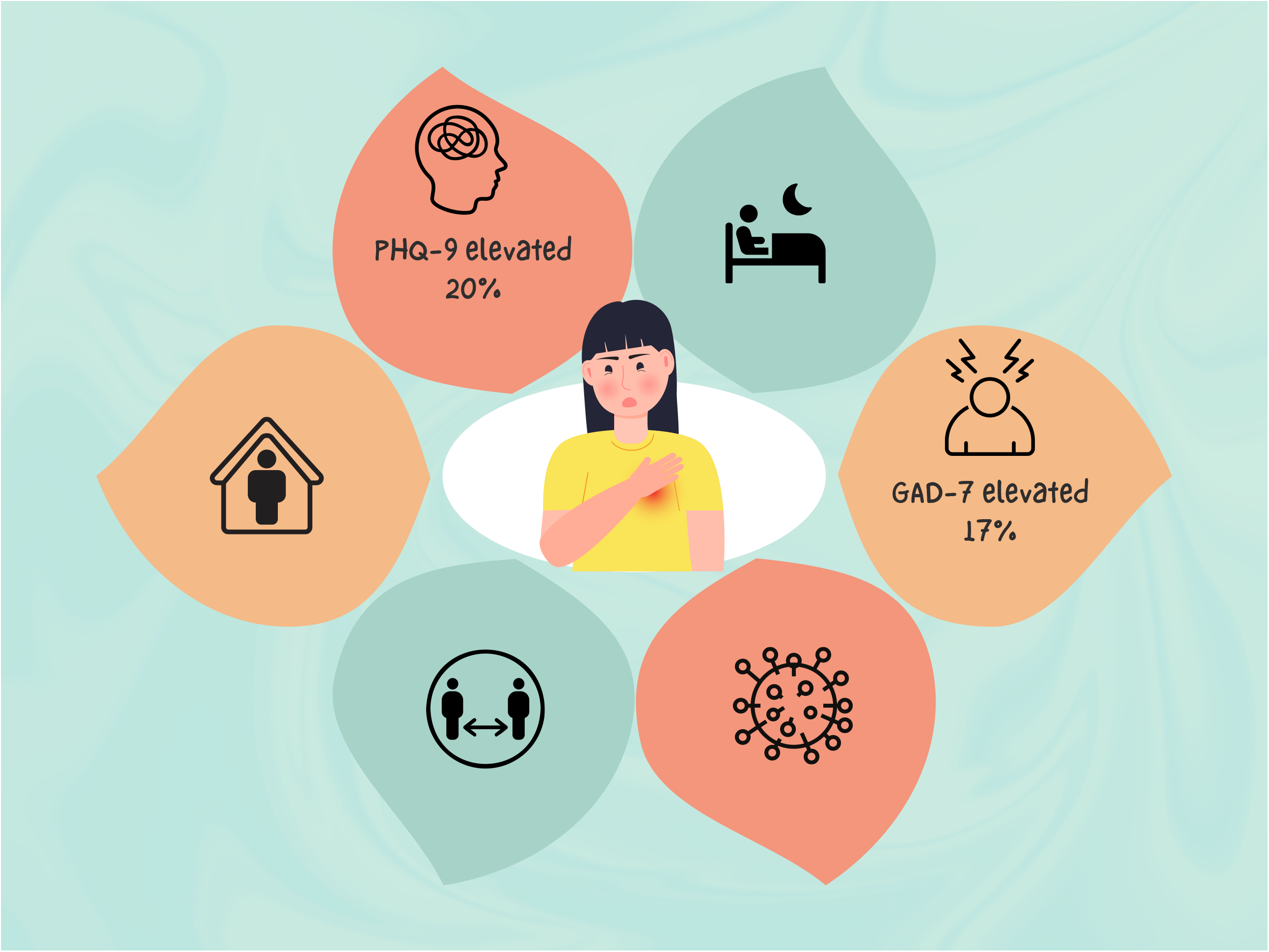
Early in the COVID-19 pandemic in China, older adults were optimistic and highly compliant with prevention and control

## REFERENCES

Beauchamp, M. K., Joshi, D., McMillan, J., Erbas Oz, U., Griffith, L. E., Basta, N. E., Kirkland, S., Wolfson, C., Raina, P., & Canadian Longitudinal Study on Aging (CLSA) Team (2022). Assessment of Functional Mobility After COVID-19 in Adults Aged 50 Years or Older in the Canadian Longitudinal Study on Aging. JAMA network open, 5(1), e2146168.

Bradley, S. M., & Rumsfeld, J. S. (2015). Depression and cardiovascular disease. Trends in cardiovascular medicine, 25(7), 614–622.

Castañeda-Babarro, A., Arbillaga-Etxarri, A., Gutiérrez-Santamaría, B., & Coca, A. (2020). Physical Activity Change during COVID-19 Confinement. International journal of environmental research and public health, 17(18), 6878.

De Pue, S., Gillebert, C., Dierckx, E., Vanderhasselt, M. A., De Raedt, R., & Van den Bussche, E. (2021). The impact of the COVID-19 pandemic on wellbeing and cognitive functioning of older adults. Scientific reports, 11(1), 4636.

Fakoya, O. A., McCorry, N. K., & Donnelly, M. (2020). Loneliness and social isolation interventions for older adults: a scoping review of reviews. BMC public health, 20(1), 129.

Fiske, A., Wetherell, J. L., & Gatz, M. (2009). Depression in older adults. Annual review of clinical psychology, 5, 363–389.

Girum, T., Lentiro, K., Geremew, M., Migora, B., Shewamare, S., & Shimbre, M. S. (2021). Optimal strategies for COVID-19 prevention from global evidence achieved through social distancing, stay at home, travel restriction and lockdown: a systematic review. Archives of public health = Archives belges de sante publique, 79(1), 150.

Hale T, Angrist N, Goldszmidt R, Kira B, Petherick A, Phillips T, Webster S, Cameron-Blake E, Hallas L, Majumdar S, Tatlow H. (2021). A global panel database of pandemic policies (Oxford COVID-19 Government Response Tracker). Nature human behaviour, 5(4), 529–538.

He, X., Li, C., Qian, J., Cui, H. (2010). Reliability and validity of a generalized anxiety scale in general hospital outpatients. Shanghai Archives of Psychiatry, 200–203.

Hunot, V., Churchill, R., Teixeira, V., Silva de Lima, M. (2007). Psychological therapies for generalised anxiety disorder. Cochrane Database of Systematic Reviews, 1, CD001848.

Izcovich, A., Ragusa, M. A., Tortosa, F., Lavena Marzio, M. A., Agnoletti, C., Bengolea, A., Ceirano, A., Espinosa, F., Saavedra, E., Sanguine, V., Tassara, A., Cid, C., Catalano, H. N., Agarwal, A., Foroutan, F., & Rada, G. (2020). Prognostic factors for severity and mortality in patients infected with COVID-19: A systematic review. PloS one, 15(11), e0241955.

Jiang, W., Sun, F., Prieto, L., Fang, Y., Gao, Y., Yue, L., Lin, X., Zhao, L., Dang, J., Qiu, J., & Li, X. (2020). Worries, strategies, and confidence of older Chinese adults during the 2019 novel coronavirus outbreak. International journal of geriatric psychiatry, 35(12), 1458–1465.

Kroenke, K., Spitzer, R. L., & Williams, J. B. (2001). The PHQ-9: validity of a brief depression severity measure. Journal of general internal medicine, 16(9), 606–613.

Lee, A., Mo, F., & Lam, L. (2020). Higher psychogeriatric admissions in COVID-19 than in severe acute respiratory syndrome. International journal of geriatric psychiatry, 35(12), 1449– 1457.

Lichtman, J. H., Froelicher, E. S., Blumenthal, J. A., Carney, R. M., Doering, L. V., Frasure-Smith, N., Freedland, K. E., Jaffe, A. S., Leifheit-Limson, E. C., Sheps, D. S., Vaccarino, V., Wulsin, L., & American Heart Association Statistics Committee of the Council on Epidemiology and Prevention and the Council on Cardiovascular and Stroke Nursing (2014). Depression as a risk factor for poor prognosis among patients with acute coronary syndrome: systematic review and recommendations: a scientific statement from the American Heart Association. Circulation, 129(12), 1350–1369.

Lorant, V., Smith, P., Van den Broeck, K., & Nicaise, P. (2021). Psychological distress associated with the COVID-19 pandemic and suppression measures during the first wave in Belgium. BMC psychiatry, 21(1), 112.

Löwe, B., Decker, O., Müller, S., Brähler, E., Schellberg, D., Herzog, W., & Herzberg, P. Y. (2008). Validation and standardization of the Generalized Anxiety Disorder Screener (GAD-7) in the general population. Medical care, 46(3), 266–274.

McGinty, E. E., Presskreischer, R., Han, H., & Barry, C. L. (2022). Trends in Psychological Distress Among US Adults During Different Phases of the COVID-19 Pandemic. JAMA network open, 5(1), e2144776.

Ping, W., Zheng, J., Niu, X., Guo, C., Zhang, J., Yang, H., & Shi, Y. (2020). Evaluation of health-related quality of life using EQ-5D in China during the COVID-19 pandemic. PloS one, 15(6), e0234850.

Robinson, E., Sutin, A. R., Daly, M., & Jones, A. (2022). A systematic review and meta-analysis of longitudinal cohort studies comparing mental health before versus during the COVID-19 pandemic in 2020. Journal of affective disorders, 296, 567–576.

Sams, N., Fisher, D. M., Mata-Greve, F., Johnson, M., Pullmann, M. D., Raue, P. J., Renn, B. N., Duffy, J., Darnell, D., Fillipo, I. G., Allred, R., Huynh, K., Friedman, E., & Areán, P. A. (2021). Understanding Psychological Distress and Protective Factors Amongst Older Adults During the COVID-19 Pandemic. The American journal of geriatric, 29(9), 881–894.

Santiago de Araújo Pio, C., Marzolini, S., Pakosh, M., & Grace, S. L. (2017). Effect of Cardiac Rehabilitation Dose on Mortality and Morbidity: A Systematic Review and Meta-regression Analysis. Mayo Clinic proceedings, 92(11), 1644–1659.

Schroeck, J. L., Ford, J., Conway, E. L., Kurtzhalts, K. E., Gee, M. E., Vollmer, K. A., & Mergenhagen, K. A. (2016). Review of Safety and Efficacy of Sleep Medicines in Older Adults. Clinical therapeutics, 38(11), 2340–2372.

Shahid, Z., Kalayanamitra, R., McClafferty, B., Kepko, D., Ramgobin, D., Patel, R., Aggarwal, C. S., Vunnam, R., Sahu, N., Bhatt, D., Jones, K., Golamari, R., & Jain, R. (2020). COVID-19 and Older Adults: What We Know. Journal of the American Geriatrics Society, 68(5), 926–929.

Tian, F., Li, H., Tian, S., Yang, J., Shao, J., & Tian, C. (2020). Psychological symptoms of ordinary Chinese citizens based on SCL-90 during the level I emergency response to COVID-19. Psychiatry research, 288, 112992.

Wang, W., Bian, Q., Zhao, Y., Li, X., Wang, W., Du, J., Zhang, G., Zhou, Q., & Zhao, M. (2014). Reliability and validity of the Chinese version of the Patient Health Questionnaire (PHQ-9) in the general population. General hospital psychiatry, 36(5), 539–544.

Wilson, K., Mottram, P.G., Sivananthan, A., Nightingale, A. (2001). Antidepressants versus placebo for the depressed elderly. Cochrane Database of Systematic Reviews, 2. CD000561.

Wong, S., Zhang, D., Sit, R., Yip, B., Chung, R. Y., Wong, C., Chan, D., Sun, W., Kwok, K. O., & Mercer, S. W. (2020). Impact of COVID-19 on loneliness, mental health, and health service utilisation: a prospective cohort study of older adults with multimorbidity in primary care. The British journal of general practice, 70(700), e817–e824.

World of Atlas (2022). https://www.worldatlas.com (last accessed 10 October 2022)

Wu, Y., Tao, Z., Qiao, Y., Chai, Y., Liu, Q., Lu, Q., Zhou, H., Li, S., Mao, J., Jiang, M., & Pu, J. (2022). Prevalence and characteristics of somatic symptom disorder in the elderly in a community-based population: a large-scale cross-sectional study in China. BMC psychiatry, 22(1), 257.

Xiong, J., Lipsitz, O., Nasri, F., Lui, L., Gill, H., Phan, L., Chen-Li, D., Iacobucci, M., Ho, R., Majeed, A., & McIntyre, R. S. (2020). Impact of COVID-19 pandemic on mental health in the general population: A systematic review. Journal of affective disorders, 277, 55–64.

Ye, Y. Y., Chen, N. K., Chen, J., Liu, J., Lin, L., Liu, Y. Z., Lang, Y., Li, X. J., Yang, X. J., & Jiang, X. J. (2016). Internet-based cognitive-behavioural therapy for insomnia (ICBT-i): a meta-analysis of randomised controlled trials. BMJ open, 6(11), e010707.

Zhang, Y., Wang, S., Ding, W., Meng, Y., Hu, H., Liu, Z., Zeng, X., Guan, Y., & Wang, M. (2020). Status and influential factors of anxiety depression and insomnia symptoms in the work resumption period of COVID-19 epidemic: A multicenter cross-sectional study. Journal of psychosomatic research, 138, 110253.

Zhao, S. Z., Wong, J., Luk, T. T., Wai, A., Lam, T. H., & Wang, M. P. (2020). Mental health crisis under COVID-19 pandemic in Hong Kong, China. International journal of infectious diseases, 100, 431–433.

Zheng, C., Shao, W., Chen, X., Zhang, B., Wang, G., & Zhang, W. (2022). Real-world effectiveness of COVID-19 vaccines: a literature review and meta-analysis. International journal of infectious diseases, 114, 252–260.

Zhuang, Q., Mao, J., Li, C., He, B. (2010). Developing of somatic self-rating scale and its reliability and validity. Chinese Journal of Behavioral Medicine and Brain Science, 9, 847–849.

